# Association of antiseizure medication with lower amyloid and tau burden

**DOI:** 10.64898/2026.06.22.26356204

**Authors:** Carolina Ferreira-Atuesta, Kai Michael Schubert, Daniela Noain, Bogdan Draganski, Marian Galovic, the Alzheimer’s Disease Neuroimaging Initiative

## Abstract

Network hyperexcitability is increasingly implicated in prodromal Alzheimer’s disease and may be suppressed by antiseizure medications (ASMs). ASMs are widely prescribed to older adults, yet whether their use relates to Alzheimer’s-disease biomarkers at the population level is unknown. In 52,537 participants in the National Alzheimer’s Coordinating Center (NACC) study, we compared cerebrospinal-fluid biomarkers, amyloid and tau positron emission tomography (PET) between ASM users and non-users using inverse-probability-of-treatment weighting with gradient-boosted propensity scores. ASM users showed directionally lower amyloid across multiple brain regions, amplifying markedly in APOE ε4 carriers (Centiloid β = −25.7, p = 0.007). All three temporal tau-PET composites were significantly lower in users (META-temporal β = −0.05, p = 0.01). The amyloid finding replicated independently in the the Alzheimer’s Disease Neuroimaging Initiative (ADNI) dataset (Centiloid β = −8.6, p = 0.01), whereas four comparator drug classes showed no amyloid signal. These convergent observational findings provide a quantitative framework for evaluating ASMs as candidate disease-modifying agents in Alzheimer’s disease.

## Background

Alzheimer’s disease is characterised by the progressive accumulation of amyloid-β plaques and neurofibrillary tau tangles, accompanied by neurodegeneration and cognitive decline.^1^ While amyloid and tau pathology disrupts neuronal and synaptic function,^2,3^ neuronal activity reciprocally shapes the production, deposition and clearance of these proteins.^4^ This two-way relationship raises the possibility that modulating neuronal activity could alter the course of amyloid and tau pathology.^5^

In preclinical models, amyloid-β is released into the interstitial fluid in an activity-dependent manner, and regional activity predicts where it is subsequently deposited or cleared ^6,7^ Amyloid-β, in turn, induces neuronal hyperactivity: soluble oligomers impair astrocytic glutamate reuptake and sustain a self-reinforcing vicious cycle of overactivation around plaques.^4,8^ Attenuating this activity restores network function and improves cognition in mouse models ^9,10^, and neuronal activity also regulates the trans-synaptic spread of tau^11–13^. Neuronal hyperactivity thus represents an early and potentially modifiable contributor to both proteinopathies.^14–16^

These findings are not limited to animal models. On sensitive electrophysiological recording, subclinical epileptiform activity is detectable in approximately 40% of patients with Alzheimer’s disease, most of whom never experience a clinical seizure.^17^ Its presence is associated with earlier progression to dementia and a two- to three-fold faster decline in MMSE score^17^. Furthermore, intracranial EEG has localised this activity to regions with elevated amyloid and tau PET signal.^18^ Hyperexcitability thus appears to be an intrinsic feature of the disease.

These observations have prompted efforts to slow pathological protein accumulation by pharmacologically reducing neuronal activity.^6,19,20^ Low-dose levetiracetam normalised hippocampal hyperactivity and improved memory in amnestic mild cognitive impairment ^21,22^ and reduced epileptiform activity in a subsequent trial. Despite this; cognitive benefits have been inconsistent and a further placebo-controlled trial was limited by recruitment.^23^ Most work has focused almost exclusively on levetiracetam, yet the medications capable of reducing neuronal activity are pharmacologically heterogeneous and may not be equivalent in their effects on protein accumulation. Consistent with this, we recently observed that older adults with epilepsy treated with sodium-channel blockers had a 27% lower risk of incident dementia than those treated with levetiracetam^24^. However, whether ASM use relates to Alzheimer’s-disease biomarkers at the population level is unknown, and there is little evidence for in vivo reduction of amyloid and tau burden in humans.

Here, we examined whether ASM exposure is associated with Alzheimer’s disease biomarkers in 52,537 older adults in the Uniform Data Set of the National Alzheimer’s Coordinating Center (NACC). ^25^ Using inverse-probability-of-treatment weighting of users and non-users, we compared a multimodal panel spanning CSF biomarkers and amyloid and tau positron-emission tomography (PET). We validated the findings in the Alzheimer’s Disease Neuroimaging Initiative (ADNI) cohort and compared them against other commonly prescribed drugs.

## Results

### Cohort and exposure

#### NACC

Of 52,537 participants enrolled across 39 NACC centers between 2005 and 2024, 3,828 had at least one cerebrospinal-fluid (CSF) or PET biomarker measurement (CSF, 2,215; amyloid PET, 1,576; tau PET, 912). Among them, 141 of those with CSF, 182 of those with amyloid PET, and 136 of those with tau PET had documented ASM-use before the measurement, forming the exposed group in each analysis ( **Extended Data table 1, Extended Data Figure 1**). ASM users and non-users were similar on age (median 69 versus 70 years), sex, education and cognitive stage, with comparable MMSE, MoCA and CDR scores (all p > 0.08), and APOE ε4 carriage was slightly lower in users (34.7% versus 40.4%; p = 0.009). The groups differed mainly in the indications for ASM use, with users more often having documented seizures (5.7% versus 1.3%), anxiety or psychiatric medication use, and a higher burden of hypertension and diabetes (each p < 0.01).

#### ADNI

Of 4,808 participants enrolled in ADNI between 2005 and 2026, 2,579 had at least one cerebrospinal-fluid (CSF) or PET biomarker measurement (CSF, 1,660; amyloid PET, 2,228; tau PET, 1,439). Among them, 76 of those with CSF, 138 of those with amyloid PET and 137 of those with tau PET had documented ASM use before the measurement, forming the exposed group in each analysis ( **Extended Data table 1, Extended Data Figure 1**). ASM users and non-users were similar in age (median 71 versus 73 years), sex, education and cognitive stage, with comparable MMSE, MoCA and CDR scores (all p > 0.48), and APOE ε4 carriage did not differ significantly (41.9% versus 45.3%; p = 0.65). As in NACC, the groups differed mainly in the indications for ASM use: users more often had documented seizures (5.4% versus 1.2%), anxiety or psychiatric medication use, and a higher burden of diabetes (14.9% versus 10.8%) (each p < 0.01), whereas hypertension was comparable between groups (50.0% versus 49.6%).

### Covariate balance and weighting precision

#### NACC

Across the multimodal panel, inverse-probability-of-treatment weighting on the 23-covariate baseline set (see Methods) reduced covariate imbalance to a mean weighted absolute standardised mean difference of 0.066 in every pooled arm. For the larger baseline-anchored outcomes, no covariate retained a weighted absolute standardised mean difference above 0.10 after weighting. In the smaller biomarker strata (CSF, amyloid PET and tau PET), residual imbalance persisted on a minority of covariate–outcome pairs after weighting and common-support trimming (47 pairs for CSF, 72 for amyloid PET, and 30 for tau PET); every residually-imbalanced covariate was carried into the outcome model as a doubly-robust adjustment (**Supplementary Table S1**).

#### ADNI

Across the multimodal panel, inverse-probability-of-treatment weighting on the 25-covariate baseline set (Methods) reduced covariate imbalance to a mean weighted absolute standardised mean difference of 0.045 across the pooled arms. Within the biomarker strata (CSF, amyloid PET and tau PET), residual imbalance remained on only a small minority of covariate-by-outcome pairs after weighting and common-support trimming (14 for CSF, 19 for amyloid PET and 4 for tau PET), and every such covariate was carried into the corresponding outcome model as a doubly-robust adjustment.

After truncation, the treated-arm effective sample size (ESS) retained 63–76% of the raw treated sample size across the headline biomarker arms in NACC (n treated = 88–131; ESS = 60–88) and 67–76% across the corresponding arms in ADNI (n treated = 47–103; ESS = 33–78). Maximum-to-median weight ratios were 1.5–2.4 in NACC and 1.5–1.7 in ADNI, indicating that no small subset of participants carried disproportionate weight. Approximately 2% of weights were clipped by truncation in every arm, and the 1st/99th-percentile cut-off recovered ∼5–10 ESS units of precision per arm without distorting the underlying weight distribution **(Supplementary Table S2)**.

### Pooled effects across the multimodal outcome panel Cerebrospinal-fluid biomarkers

The p-tau/amyloid-β42 ratio was significantly lower in users (β = −0.217; 95% CI −0.385 to −0.050; p = 0.011; Cohen’s d = −0.23; N = 1,312 of which users = 100) (**Figure 1A**). CSF amyloid-β42 was directionally higher in users than in non-users, consistent with lower amyloid burden, although the difference did not reach significance (β = +0.078; 95% CI −0.062 to +0.218; p = 0.274; Cohen’s d = +0.11; N = 1,141, of which users = 108). CSF p-tau181 (β = −0.074; 95% CI −0.242 to +0.094; p = 0.387; Cohen’s d = −0.11; N = 1,186, of which users = 102), total tau (β = +0.076; 95% CI −0.189 to +0.341; p = 0.574; Cohen’s d = +0.07; N = 1,121, of which users = 100) and the total-tau/amyloid-β42 ratio (β = +0.008; 95% CI −0.247 to +0.262; p = 0.954; Cohen’s d = +0.01; N = 1,121, of which users = 100) did not differ at the cross-sectional level (**Figure 1A**).

**Figure 1.**
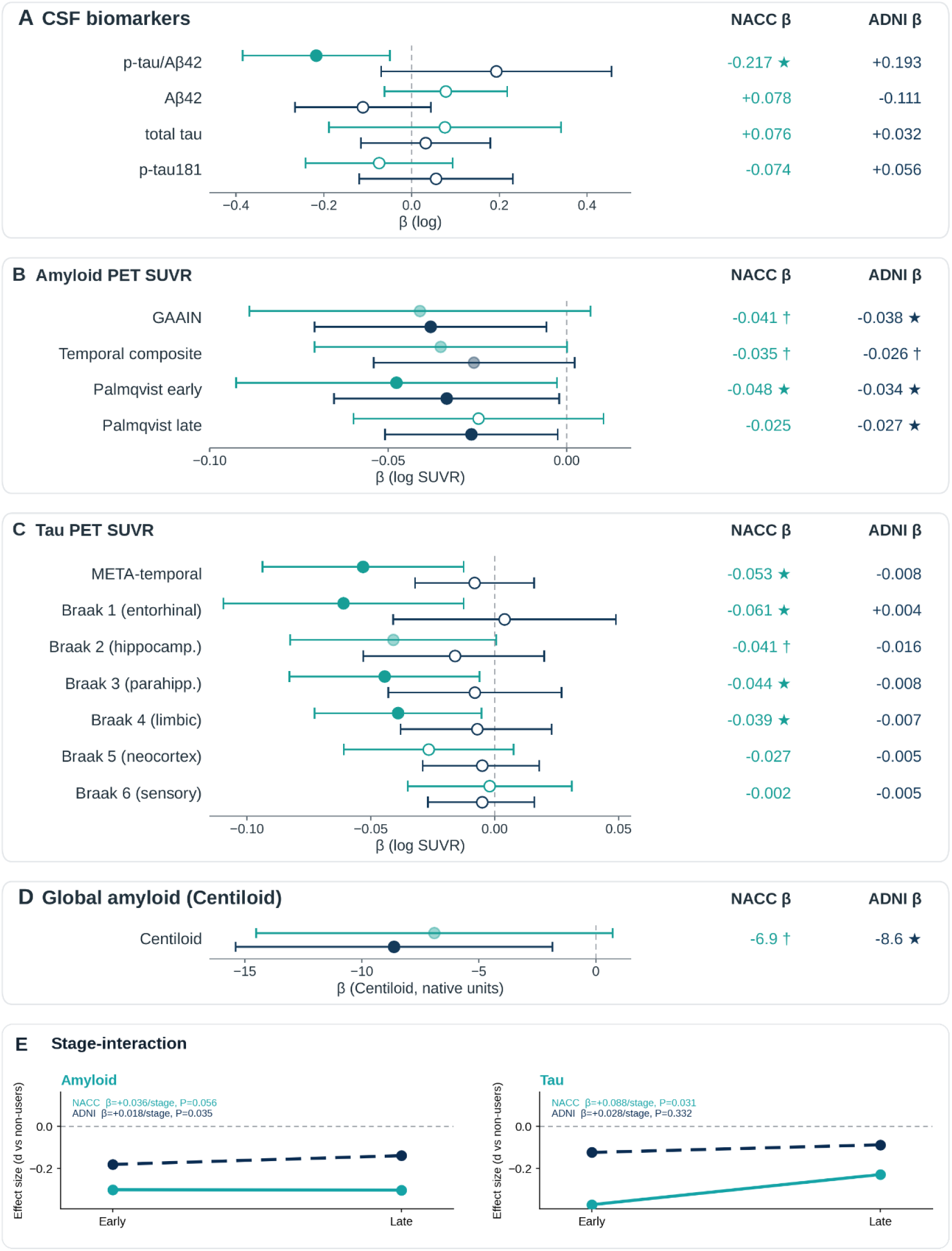
Effect of baseline ASM use on cross-sectional Alzheimer’s-disease biomarkers in NACC (teal) and ADNI (navy), from doubly-robust inverse-probability-weighted least squares. (a) CSF biomarkers; (b) amyloid-PET SUVR composites; (c) tau PET (META-temporal and Braak stages); (d) global amyloid (Centiloid); β with 95% confidence-interval whiskers (filled, p < 0.05; faded, p < 0.10; open, not significant). (e) ASM effect size (Cohen’s d, users vs non-users) across disease stage by cohort (NACC solid, ADNI dashed); the effect is largest at early disease stage and attenuates toward late stage.

### Positron-emission tomography

#### Amyloid

On amyloid PET, the Global Alzheimer’s Association Interactive Network (GAAIN) summary SUVR (β = −0.0411; 95% CI −0.0889 to +0.0067; p = 0.092; Cohen’s d = −0.15; N = 1,393, of which users = 137) and the Centiloid value (β = −6.90; 95% CI −14.52 to +0.72; p = 0.076; Cohen’s d = −0.16; N = 1,051, of which users = 126) were both directionally lower in users (**Figure 1B,D**). Bilateral hippocampal and entorhinal SUVR showed the same direction. All 17 cortical amyloid composites pointed in the negative, lower-amyloid direction (**Supplementary Table S3**). These findings were replicated in direction, magnitude and statistical significance in ADNI: ASM users had significantly lower amyloid Centiloid (β = −8.64, 95% CI −15.48 to −1.79, p = 0.013), lower summary SUVR (β = −0.040, 95% CI −0.073 to −0.007, p = 0.019), and lower hippocampal SUVR (β = −0.013, 95% CI −0.025 to −0.002, p = 0.022) (**Figure 1B**).

The amyloid signal amplified sharply in APOE ε4 carriers (**Figure 2B**), in whom the Centiloid reduction reached β = −25.67 (95% CI −44.25 to −7.10; p = 0.007; Cohen’s d = −0.61; N = 207, of which users = 17) and GAAIN summary SUVR reduction β = −0.113 (95% CI −0.200 to −0.025; p = 0.012; Cohen’s d = −0.42; N = 290, of which users = 27). By contrast, in APOE ε4 non-carriers the global amyloid signal was essentially absent (Centiloid β = −0.79; 95% CI −12.23 to +10.66; p = 0.893; Cohen’s d = −0.02; N = 456, of which users = 73; GAAIN summary SUVR β = −0.033; 95% CI −0.100 to +0.033; p = 0.322; Cohen’s d = −0.12; N = 574, of which users = 65). This APOE4-amplification pattern replicated independently in the ADNI cohort (carrier Centiloid β = −19.7, p = 0.0003; overall β = −8.64, p = 0.013) (**Figure 2B, D**).

**Figure 2.**
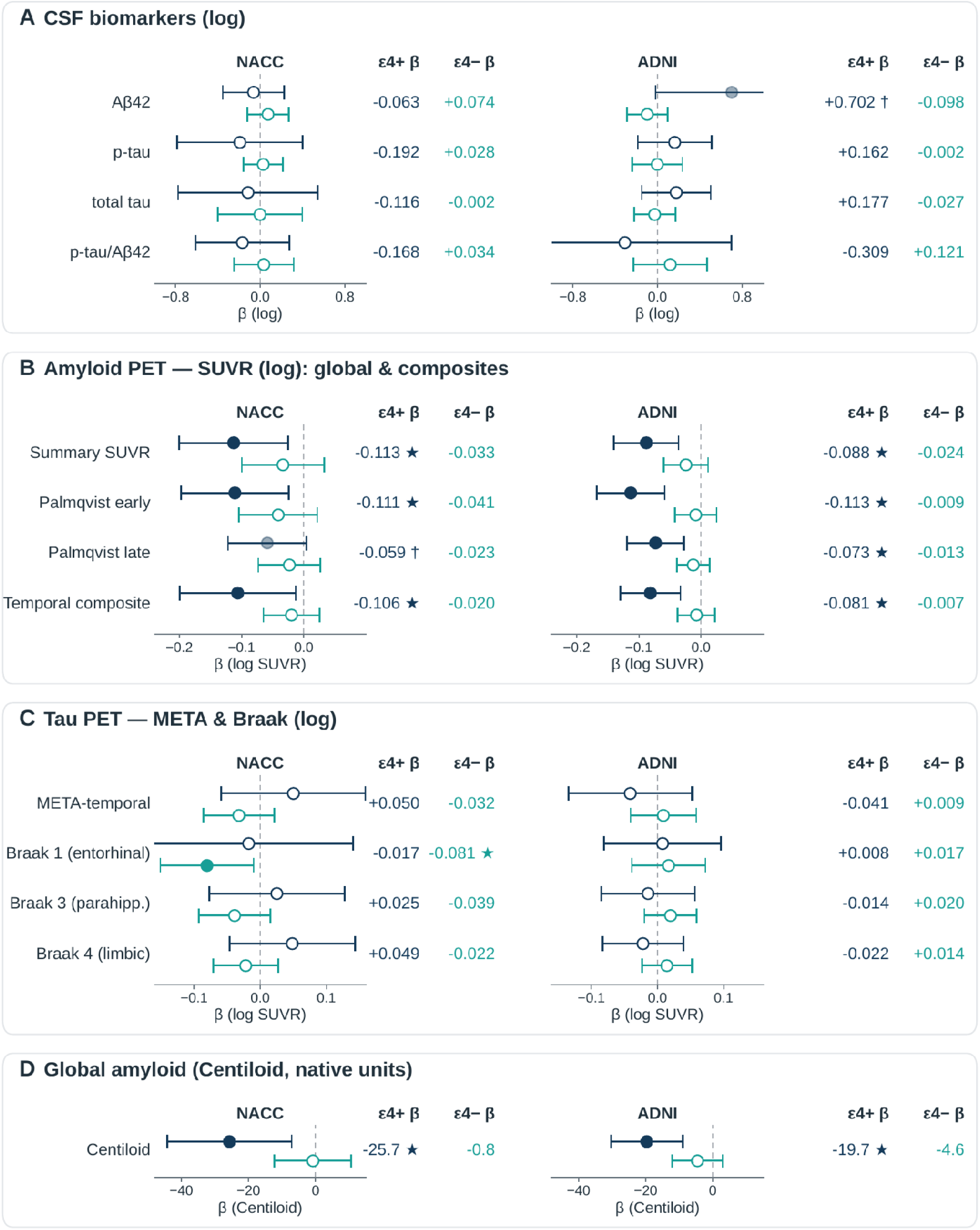
APOE ε4 modification of the ASM–biomarker association. ε4 carriers (navy; heterozygous + homozygous) versus non-carriers (teal; no ε4), with non-carriers as the common reference in both NACC and ADNI. Each outcome is one row (NACC left, ADNI right); β with exact 95% confidence intervals (filled, p < 0.05; faded, p < 0.10; open, not significant). Panels by scale: (a) CSF; (b) amyloid SUVR; (c) tau; (d) Centiloid. ADNI CSF carrier n is small (7–11 users); wide confidence intervals are clipped at the axis.

There was a positive significant interaction between ASM use and accumulation stage, indicated by the trend test⁸ (β = +0.036, p = 0.056; N = 1,393) (**Figure 1E**). This is consistent with an early-stage-anchored amyloid signal that is largest in early-accumulating regions. This interaction was directionally concordant in the ADNI replication (β = +0.0185; p = 0.035; N = 2,047) (**Figure 1E**).

#### Tau

On tau PET, the bilateral META-temporal composite was significantly lower in users (β = −0.0531; 95% CI −0.0937 to −0.0125; p = 0.010; Cohen’s d = −0.23; N = 678, of which users = 84) (**Figure 1C**), as were the medial-temporal (β = −0.0531; 95% CI −0.0954 to −0.0108; p = 0.014; N = 678, of which users = 84) and lateral-temporal (β = −0.0445; 95% CI −0.0826 to −0.0063; p = 0.022; N = 678, of which users = 84) composites. The Braak ordinal-stage analysis showed an early-stage-concentrated reduction, strongest at Braak 1 (entorhinal; β = −0.0610; 95% CI −0.1095 to −0.0126; p = 0.014), with a significant positive ASM use × stage interaction (β = +0.0109; 95% CI +0.0006 to +0.0211; p = 0.038; N = 678) indicating attenuation across later Braak stages (**Figure 1E**). APOE ε4-stratified tau analyses were limited by small per-stratum samples (carrier META-temporal β = +0.050; p = 0.366; Cohen’s d = +0.21; N = 98, of which users = 7; non-carrier β = −0.032; p = 0.236; Cohen’s d = −0.14; N = 258, of which users = 23), with the Braak 1 reduction significant only in non-carriers (β = −0.081; p = 0.025) (**Figure 2C**). ADNI tau-PET results were null at the cohort level (META-temporal β = −0.008, p = 0.509; **Supplementary Table S4**).

At the voxel map level, multiple amyloid PET clusters survived cluster-extent FWE correction at p < 0.05 in both hemispheres (left p_FWE = 0.040, right p_FWE = 0.033) (**Figure 3A**) whereas tau-PET cluster-permutation maps did not yield supra-threshold clusters despite the per-region cross-sectional regression being significant for the meta-temporal composites. The amyloid-PET surface signal replicated in ADNI under the time-anchored canonical, where bilateral amyloid clusters survived correction (left p_FWE = 0.038, right p_FWE = 0.038) (**Figure 3A**). The implicated regions overlapped the neocortical association and default-mode territory in which amyloid typically accumulates earliest in Alzheimer’s disease, including orbitofrontal and cingulate cortex. A shared core spanning these regions was common to both cohorts, while NACC additionally extended into the mesial temporal cortex that did not survive correction in ADNI. **Supplementary Table S5** contains the list of the significant ROIs and their effect sizes.

**Figure 3.**
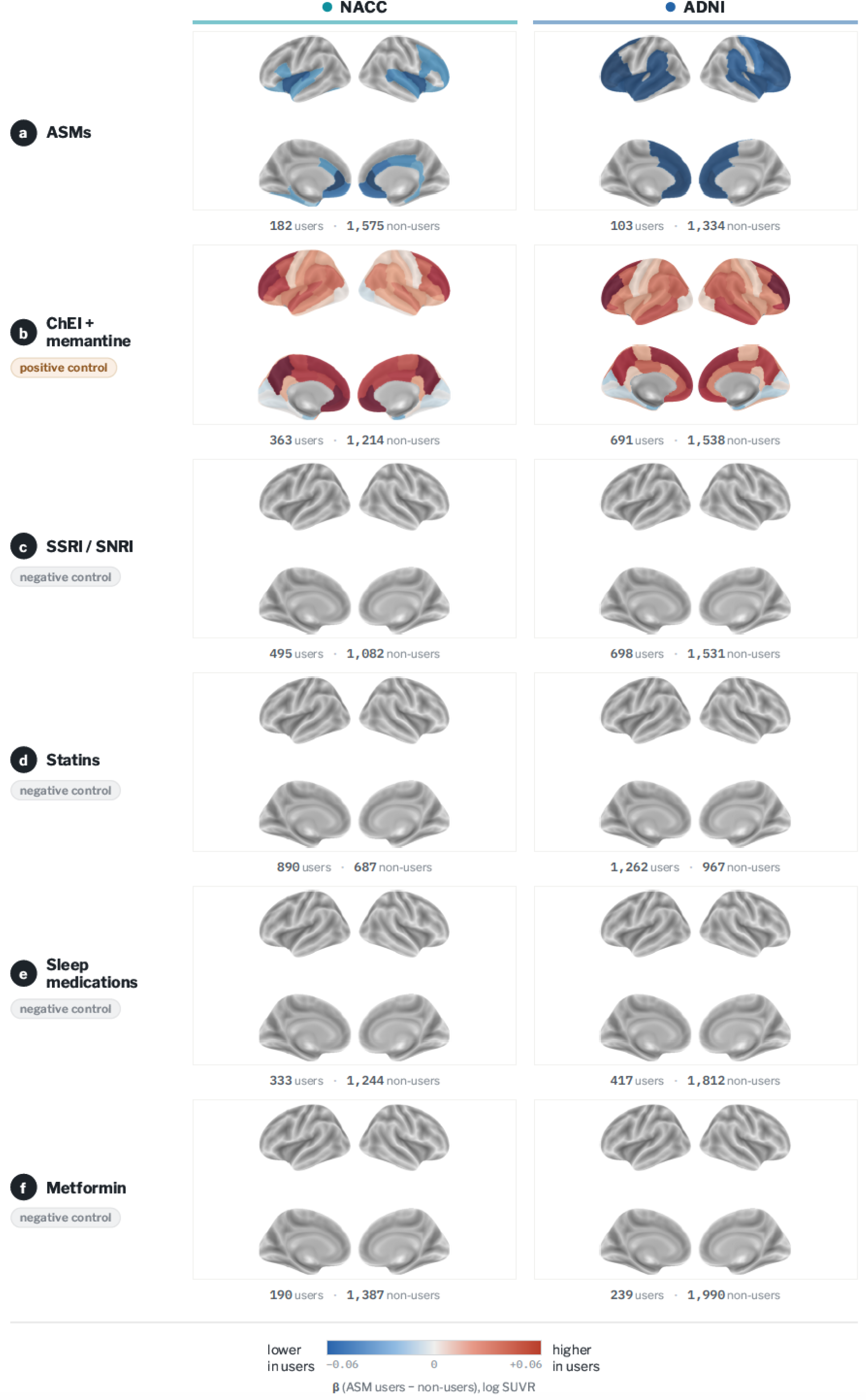
Maps of the amyloid-PET SUVR difference between users and non-users for NACC and ADNI. Colour encodes the β coefficient (ASM users − non-users, log SUVR); blue indicates lower amyloid burden in users. Regions shown survive a cluster-forming threshold p < 0.05 with family-wise-error control by 2,000 permutations (p_FWE).

### Sensitivity analyses

#### Monotherapy versus polytherapy

Within the monotherapy stratum, biomarker trends mirrored those of the primary analysis but remained below the threshold for statistical significance (CSF p-tau/Aβ42 β = −0.096, p = 0.355, Cohen’s d = −0.10; Centiloid β = −3.74, p = 0.417, Cohen’s d = −0.09; META-temporal tau β = −0.035, p = 0.080, Cohen’s d = −0.15; **Supplementary Figure S1**). Conversely, polytherapy was associated with substantially more robust reductions: users of multiple concurrent agents exhibited significantly lower CSF p-tau/Aβ42 ratios (β = −0.455, p < 0.001, Cohen’s d = −0.49) and META-temporal tau burden (β = −0.150, p < 0.001, Cohen’s d = −0.64). Cortical amyloid composites were similarly reduced (GAAIN β = −0.101, p = 0.029, Cohen’s d = −0.38; temporal β = −0.073, p = 0.048), while the global Centiloid reduction remained directional (β = −10.41, p = 0.291, Cohen’s d = −0.25), constrained by the smaller sample size of the polytherapy group (**Supplementary Figure S1**).

#### Exposure at baseline

To further mitigate potential bias and isolate the effects of sustained treatment, we restricted the cohort to participants with baseline-documented ASM use. By anchoring exposure to the initial visit, this approach focuses on prevalent users with a longer duration of medication exposure while simultaneously minimizing immortal-time bias. This baseline-anchored exposure was associated with more pronounced reductions across the headline markers (Centiloid β = −10.99, p = 0.041, Cohen’s d = −0.26; META-temporal tau β = −0.056, p = 0.048, Cohen’s d = −0.24; **Supplementary Figure S2**). While effects on regional amyloid PET composites (GAAIN summary β = −0.039, Cohen’s d = −0.15; temporal β = −0.036; both p > 0.05) and CSF analytes (p-tau/Aβ42 ratio β = −0.155, Cohen’s d = −0.17; Aβ42 β = +0.073, Cohen’s d = +0.10; both p > 0.05) remained directionally consistent, they did not reach statistical significance (**Supplementary Figure S2)**. This pattern of results is consistent with a cumulative-exposure relationship in which chronic suppression of neuronal activity has its most detectable impact on global amyloid burden and downstream tau pathology.

#### Drug-class specificity of the amyloid-PET signal

To test whether the signal was specific to ASMs, we repeated the per-region IPW surface analysis in both cohorts for five comparator drug classes: metformin, serotonergic antidepressants (SSRIs/SNRIs), statins, memantine, and sleep medications, under an identical pipeline (23-covariate set with at-or-before anchoring) and exposure definition (ever-use at or before outcome). Memantine was used as a positive control, since it is prescribed to patients with established Alzheimer’s disease. Of the five comparators, only memantine produced positive supra-threshold clusters that survived FWE05 (p = 5 × 10⁻⁴ each). Metformin, SSRIs/SNRIs, statins, and sleep medications were null or borderline on amyloid (best cluster p ≥ 0.14). The same panel on tau PET confirmed the memantine positive control (p = 0.020 and 0.023 bilaterally) and the absence of any specific signal from the other comparators (**Figure 3B-F, Supplementary Table S6**).

## Discussion

Across two independent cohorts, ASM use was associated with lower amyloid and tau markers of Alzheimer’s pathology, in what is, to our knowledge, the largest analysis of ASM exposure and Alzheimer’s-disease biomarkers to date. The amyloid signal was concentrated in APOE ε4 carriers across both cohorts, and a pre-specified comparator-drug panel yielded no protective amyloid signal in either cohort, confirming the specificity of the findings.

The regional and stage-dependent structure of these associations aligns with the established activity-dependence of amyloid and tau dynamics. Interstitial concentrations of amyloid-β rise with neuronal firing and fall when synaptic activity is suppressed ^6^, and the level of regional neuronal activity determines where amyloid-β subsequently aggregates.^7^ The cortical regions that are most metabolically active in the resting brain (posterior cingulate, precuneus and lateral temporoparietal association cortex) are also those in which amyloid is deposited earliest.^26^ Tau is governed by the same principle: its secretion is stimulated by neuronal activity^11^, and heightened activity accelerates ^27,28^ trans-synaptic tau propagation and pathology in vivo.^11^ Because network hyperexcitability is an early and modifiable feature of the disease an exposure that lowers excitability would be expected to act preferentially on these early-involved, high-activity regions rather than uniformly across lower activity regions. This is the pattern we observed: the amyloid signal localized to temporal, cingulate and insular cortex, and the stage-interaction analyses showed that attenuation was greatest in early-stage regions and diminished across successively later stages, with the largest single-region tau reduction in Braak I.

The associations were markedly stronger in APOE ε4 carriers. In carriers, ASM exposure was associated with substantially lower amyloid burden and concordant reductions in the early-deposition composites whereas in non-carriers the same contrasts were small and non-significant. Recent work shows that neuronal APOE4 drives early, region-specific hippocampal network hyperexcitability that precedes pathology and predicts later cognitive deficits, and that this phenotype is abolished by selective removal of neuronal APOE4^29^; ε4 also impairs hilar GABAergic interneurons and shifts the excitation–inhibition balance toward disinhibition.^30^ A brain already biased toward hyperexcitability by ε4 would be expected to derive a larger benefit from an exposure that lowers neuronal activity, which is the direction we observed.

The comparator-drug panel reinforced this interpretation. Of the five classes tested, the four with indications distinct from those of ASMs, namely metformin, serotonergic antidepressants (SSRIs/SNRIs), statins and sleep medications, produced no amyloid signal in either cohort, with best cluster-level p-values ≥ 0.14, indicating that the association is not a generic feature of chronic medication use in this population.

Finally, the association was larger when exposure was restricted to sustained, baseline-documented use than in the pooled analysis, consistent with a cumulative-exposure relationship in which greater suppression of neuronal excitability produces a correspondingly greater reduction in activity-dependent pathology.

The clinical implications of our results are non-trivial. The ASMs are off-patent, widely prescribed, generally well-tolerated, and have a decades-long real-world safety record in older adults. The biological mechanisms have independently been demonstrated in transgenic models to affect network hyperexcitability, tau seeding, and amyloid clearance ²³⁵⁶⁷, and the population-level ever-user signal is consistent with these mechanisms. These findings provide a quantitative observational basis for prioritising one or more of these widely-available agents for prospective disease-modification trials in pre-clinical or prodromal Alzheimer’s disease.

Our study has several strengths. To our knowledge it is the largest analysis of ASM exposure and Alzheimer’s-disease biomarkers to date, drawing on 52,537 participants across 39 Alzheimer’s Disease Research Centers together with an independent replication cohort. Rather than collapsing the disease into a single endpoint, we assembled a multimodal outcome panel spanning CSF, amyloid PET and tau PET, which allowed the same exposure to be interrogated against molecularly distinct markers and revealed a convergent pattern that no single modality could establish on its own. The analytic design was built specifically to counter the two dominant threats to validity in pharmaco-epidemiology: Confounding by indication was addressed through gradient-boosted propensity-score estimation, common-support trimming and a doubly-robust weighted-outcome specification in which any residually-imbalanced covariate re-entered the outcome model while reverse causation was addressed by anchoring exposure to documentation at or before each outcome and by the baseline-only sensitivity analysis, which additionally showed larger effects under sustained exposure, consistent with a biological rather than a confounded signal.

Per-outcome weighting preserved the available sample within each biomarker stratum rather than discarding it through hard matching. The primary biomarker outcomes and the tau and amyloid progression tests were pre-specified, limiting the scope for selective reporting, and the central findings reproduced in ADNI, a cohort with independent recruitment, covariate structure and PET-processing pipelines. Finally, the pre-specified comparator-drug panel and the memantine positive control provided an internal calibration of the pipeline that few observational biomarker analyses include.

Several limitations temper these conclusions. First, exposure is asserted by chart review: the NACC current-medication form records only whether a drug appears on a participant’s medication list at a given visit, without dose, frequency, duration or adherence, so a single low-dose prescription cannot be distinguished from years of sustained high-dose use, and any exposure preceding cohort entry is not captured, a misclassification that would, if anything, bias a true effect toward the null. Second, the design is observational, and although weighting and doubly-robust adjustment remove measured confounding, residual confounding by unmeasured factors cannot be excluded; in particular, the indications for which these agents are prescribed (neuropathic pain, anxiety, mood symptoms, seizures) may themselves index biological phenotypes correlated with Alzheimer’s pathology, and the comparator and positive-control analyses bound but do not eliminate this possibility. Third, the NACC sample is a convenience cohort of adults presenting to Alzheimer’s Disease Research Centers, enriched for cognitive concern and for white participants relative to the United States older-adult population, which limits generalisability; replication in ADNI, a demographically similar research cohort, mitigates but does not resolve this. Fourth, once exposure is anchored before each outcome the biomarker strata are modest, the APOE-stratified and tau-PET analyses rest on small per-stratum samples, and the amyloid- and tau-PET longitudinal accumulation analyses were under-powered (n = 6 and n = 6 users with ≥ 2 scans, respectively). Fifth, because more than 500 hypothesis tests are reported we did not impose a single experiment-wide correction, and per-drug and per-region findings are therefore treated as hypothesis-generating throughout; we note, however, that the headline findings (the CSF p-tau/amyloid-β42 ratio, the temporal-lobe tau composites, and the amyloid Palmqvist and Braak progression interactions) survive Benjamini-Hochberg correction within their outcome category. Together these constraints place the present results at the level of a population-scale association that motivates, rather than substitutes for, prospective interventional testing.

Taken together, these findings position ASM exposure as a population-level signal of reduced cortical amyloid and reduced temporal-lobe tau accumulation, consistent across CSF, amyloid-PET and tau-PET modalities, robust to multiple IPW specifications, replicated in an independent cohort (ADNI), and supported by mechanistic precedent in transgenic models. Because the implicated drugs are inexpensive, off-patent, decades-old and broadly tolerated in older adults, they meet the operational definition of drug-repurposing candidates with a substantially lower clinical-development risk than novel disease-modifying agents. We recommend that the highest-priority next step is a placebo-controlled randomised trial of low-dose chronic exposure to one or more of these agents in pre-clinical or prodromal Alzheimer’s disease, with amyloid-PET, the Palmqvist early-stage composite and the CSF p-tau/Aβ42 ratio as biomarker endpoints. The observational evidence presented here, now further supported by external replication in ADNI, provides a quantitative basis for such a trial.

## Methods

### Cohort and ethics

We used the Uniform Data Set of the National Alzheimer’s Coordinating Center (NACC data freeze v67, June 2024)^25^, comprising 52,537 unique participants enrolled across 39 federally funded United States Alzheimer’s Disease Research Centers between September 2005 and June 2024. Data collection is approved at each contributing centre by the local institutional review board, and all participants provide written informed consent. Secondary analyses of de-identified NACC data are exempt from additional institutional-review-board review at our institution. We additionally used NACC neuroimaging linkage files for amyloid PET (regional SUVRs and centiloid values), tau PET (regional SUVRs from flortaucipir and MK-6240).

Data used in the preparation of this article were obtained from the Alzheimer’s Disease Neuroimaging Initiative (ADNI) database (adni.loni.usc.edu). The ADNI was launched in 2003 as a public-private partnership, led by Principal Investigator Michael W. Weiner, MD. The primary goal of ADNI has been to test whether serial magnetic resonance imaging (MRI), positron emission tomography (PET), other biological markers, and clinical and neuropsychological assessment can be combined to measure the progression of mild cognitive impairment (MCI) and early Alzheimer’s disease (AD).

### Data cleaning and variable derivation

NACC encodes missingness, refusal and “not administered” with out-of-range integer codes; all instances of −4, 99, 8888 and 9999 in numeric columns were set to missing, and for a defined set of diagnostic flags code 8 was additionally set to missing.

Demographic and clinical covariates were derived as follows: age at first visit; sex; education in years, with missing values imputed by within-participant mode; apolipoprotein-E ε4 carrier status with a flag for unascertained genotype; race and ethnicity; and twenty comorbidity and binary diagnosis flags (stroke, Parkinson’s disease, traumatic brain injury, diabetes, hypertension, hypercholesterolaemia, sleep apnoea, atrial fibrillation, myocardial infarction, congestive heart failure, psychiatric disorder, thyroid disease, vitamin-B12 deficiency, anxiety, bipolar disorder, post-traumatic stress disorder, MCI, dementia, depression and epilepsy, the last defined as a recorded epilepsy diagnosis or as documented seizures together with any ASM use).

### Biomarker distributions and quality control

Before weighting, we examined the distribution of every continuous biomarker. CSF amyloid-β42, p-tau181 and total tau were right-skewed and approximately log-normal and were natural-log-transformed for analysis; values outside the plausible assay range of 0 to 10,000 pg ml⁻¹ were set to missing.

### Antiseizure medication ascertainment

NACC records a participant’s current-medication list at each visit across forty free-text drug columns(DRUG1–40). At each visit, every drug field was searched for each canonical ASM name and its well-established brand aliases. Fourteen agents with at least twenty documented users in the full cohort were retained: gabapentin, clonazepam, levetiracetam, pregabalin, lamotrigine, topiramate, primidone, phenytoin, carbamazepine, oxcarbazepine, valproic acid, zonisamide, phenobarbital and lacosamide. For each participant we computed, per visit, whether any medication was present and the count of distinct medications; and, per participant, an ever-use indicator, the number of visits with documented use, the maximum number of concurrent medications at any visit, and the earliest visit date reporting any medication.

Full ADNI cohort definition, data cleaning and variable derivation are described in the Supplementary Methods.

### Exposure criteria

Primary exposure was ever-use of any ASM with documentation at a visit dated on or before the outcome-measurement date. For the sensitivity analyses, different exposure criteria were applied. To address reverse causation, for every analytic outcome we defined a per-outcome anchor date and required that the exposure variable be documented at a visit dated on or before that anchor. Per-participant filtered exposure features were computed separately for each outcome category by restricting the visit set to visits dated on or before the relevant anchor.

### Outcome construction

Cerebrospinal-fluid biomarkers were taken from the earliest lumbar puncture per participant, with values outside the interval (0, 10,000] pg ml⁻¹ set to missing and the p-tau/amyloid-β42 and total-tau/amyloid-β42 ratios computed from the corresponding raw values.

Amyloid and tau PET outcomes were taken from the earliest scan of each modality per participant; bilateral hippocampus, entorhinal and precuneus SUVR were computed as means of the left and right parcels; Centiloid values were taken from the linkage file; 17 amyloid composites and the tau Braak^31^ and lobar regions were computed as means across their constituent FreeSurfer parcels^32^. For the pre-computed global amyloid metrics we additionally analysed the GAAIN Summary SUVR and Centiloid value, all taken directly from the NACC amyloid-PET linkage files and pre-harmonised across tracers (florbetaben, florbetapir, flutemetamol and NAV4694). The pre-specified tau progressive-accumulation test was a weighted long-format regression of log(tau SUVR) on the user-by-Braak-stage interaction; the analogous pre-specified amyloid early-versus-late test contrasted Palmqvist early and late composites⁸

### Statistical analysis

For every analytic outcome we estimated the effect of ASM exposure by inverse-probability-of-treatment weighting (IPW)²⁰–²². Per-outcome propensity scores were fitted with gradient boosting (scikit-learn, GradientBoostingClassifier: 200 trees, depth 3, learning rate 0.05, subsample 0.9, random seed 42) on a 23-covariate baseline set comprising six demographic covariates (age, sex, education, Black race, Asian race, Hispanic ethnicity), two genetic covariates (APOE ε4 carrier and a missing-genotype indicator), four neurological comorbidities (epilepsy, stroke, TBI, Parkinson’s disease), five cardiometabolic covariates (diabetes, hypertension, hypercholesterolaemia, atrial fibrillation, and an aggregate cardiac-history flag medhist_card = AFib/MI/CHF), three psychiatric flags (depression, anxiety, other psychiatric disorder), one endocrine flag (thyroid disease), two baseline-neuropsychiatry covariates (baseline MMSE, median-imputed when missing, and an MMSE-missingness indicator) and At-scan age, dementia and MCI indicators were added inside the cross-sectional outcome model. Every comorbidity and any-ASM exposure flag was anchored on or before the modality-specific outcome date (the lumbar-puncture date for CSF, the scan date for PET). Subjects whose propensity score fell outside the overlap of treated and untreated distributions were trimmed. Stabilised IPW weights (w_t = π/PS; w_c = (1−π)/(1−PS)) were truncated at the 1st/99th percentiles. To quantify the precision cost of weighting, we computed the Kish effective sample size (ESS = (Σw)² / Σw²) separately in the treated and control arms, and summarised the weight distribution using the maximum-to-median weight ratio and the proportion of weights clipped by truncation. Cross-sectional outcome models were weighted least squares with HC1 sandwich standard errors, log-transformed for positive-valued biomarkers (CSF analytes, SUVR composites and ratios, GAAIN summary SUVR) and on the native scale for Centiloid; PET tracer (florbetaben / florbetapir / flutemetamol / NAV4694 for amyloid; flortaucipir / MK-6240 for tau) was added for PET arms. Trend models (the Palmqvist early-vs-late amyloid composites and the Braak 1–6 ordinal tau stages) used long-format weighted regression with the ASM use × stage interaction as the focal estimand and participant-clustered standard errors. APOE ε4-stratified analyses excluded participants with unknown APOE genotype and refitted the propensity model within each stratum after dropping the constant APOE covariates. A pre-specified per-region IPW + cluster-extent FWE pipeline was applied to amyloid and tau PET, with the focal effect being the per-region weighted-OLS coefficient for ASM use; clusters were formed at a signed threshold of p < 0.05 on the within-hemisphere Desikan–Killiany adjacency graph and assessed against a null distribution generated by 2,000 permutations of the exposure label within the weighted cohort.

External replication was performed in the Alzheimer’s Disease Neuroimaging Initiative (ADNI) cohort (n = 3,680; 443 ASM ever-users) using the same IPW approach with a 30-covariate time-anchored covariate set additionally including the five medication-class indication proxies.

All analyses are implemented in Python (scikit-learn 1.x, statsmodels 0.14)²⁴.

### Model specifications

#### Model

For each outcome the probability of ASM exposure was estimated by gradient boosting, e(X) = P(any-ASM = 1 | X), with X comprising 23 baseline covariates: age at baseline; sex; education (years); APOE ε4-carrier status and an APOE-missing indicator; Black race; Asian race; Hispanic ethnicity; epilepsy; stroke; traumatic brain injury; Parkinson’s disease; diabetes; hypertension; hypercholesterolaemia; atrial fibrillation; depression; anxiety; any psychiatric disorder; thyroid disease; an aggregate cardiac-history flag; an osteoporosis-treatment proxy; and the earliest baseline Mini-Mental State Examination score (median-imputed) with an MMSE-missing indicator. For cross-sectional biomarker outcomes, at-scan mild-cognitive-impairment and at-scan dementia indicators were added to X.

#### Weights

Stabilized inverse-probability-of-treatment weights were w_i = π / e(X_i) for users and w_i = (1 − π) / (1 − e(X_i)) for non-users, where π is the marginal exposure prevalence in the common-support-trimmed cohort; weights were truncated at the 1st and 99th percentiles.

#### Cross-sectional outcome model

Each outcome was fitted by weighted least squares with HC1 robust standard errors: g(Y_i) = β₀ + β₁·any-ASM_i + θ′X_i (+ tracer) + ε_i, weighted by w_i, where g(·) is the natural logarithm for positive-valued biomarkers (CSF analytes, SUVR composites and ratios) and the identity link for Centiloid; tracer was added for PET arms. The same covariate vector X entered the outcome model (a doubly-robust specification), and the reported ASM effect was β₁.

#### Progression-trend models (Braak, Palmqvist)

Stage-dependence was tested in long format by weighted linear regression with participant-clustered robust standard errors: log(SUVR_ij) = β₀ + β₁·any-ASM_i + β₂·stage_j + β₃·(any-ASM_i × stage_j) + θ′X_i + ε_ij, weighted by w_i, where stage_j is the Braak stage (1 to 6) or the Palmqvist early-versus-late contrast for region j of participant i, adjusted for at-scan age, tracer and at-scan cognitive status. The interaction β₃ is the focal estimand; clustering on participant identifier accounts for the within-participant correlation across regions and stages.

#### Outcomes

The same weighted-least-squares specification was applied across the outcome panel: CSF analytes and ratios (natural-log scale), and amyloid- and tau-PET regional and composite SUVRs and the Centiloid value (log scale for SUVRs), each regressed on the any-ASM indicator with adjustment for at-scan cognitive status, at-scan age, sex, education, PET tracer (PET outcomes) and any baseline covariate left unbalanced after weighting (doubly-robust).

### Sensitivity analyses

#### Monotherapy vs polytherapy

Participants were classified by the maximum number of concurrent ASMs at any qualifying visit (monotherapy = 1; polytherapy ≥ 2); each compared with non-users under the per-arm IPW pipeline with users of the other arm excluded from the control pool.

#### Exposure at baseline

Exposure at baseline was defined as any document ASM at the baseline visit (NACCVNUM = 1); all outcomes re-estimated under the per-arm IPW pipeline with later-onset ever-users excluded. All sensitivity analyses use the same gradient-boosted propensity model on the 23-covariate IPW set, common-support trimming, stabilised IPW weights truncated at the 1st/99th percentiles, and weighted least squares with HC1-robust standard errors for the outcome model.

#### Drug-class specificity

To assess the specificity of the amyloid-PET surface signal, the per-region inverse-probability-weighted cluster-permutation analysis was repeated in the NACC and ADNI amyloid-PET cohorts with the focal exposure replaced by five comparator CNS-active or metabolic drug classes: metformin; serotonergic antidepressants (SSRIs and SNRIs); statins; cholinesterase inhibitors and memantine; and sleep medications (Z-drugs, dual orexin-receptor antagonists, melatonin-pathway agents, and off-label sedating antidepressants). The complete list of generic and brand-name agents searched for each class is provided in **Supplementary Table S7**. Exposure for every class was ever-use at or before the first amyloid-PET scan. For each cohort × class, the 23-covariate manuscript propensity model was refit (gradient-boosted classifier, 200 trees, depth 3, learning rate 0.05), followed by common-support trimming and stabilised weights truncated at the 1st and 99th percentiles; per-region weighted least-squares models of log(SUVR) were adjusted for at-scan cognitive status, at-scan age, sex, education, PET tracer, and any residually-imbalanced covariates (doubly-robust). Cluster-extent family-wise-error correction used 2,000 permutations of the exposure label within the weighted cohort.

## Data availability

NACC data are available to qualified investigators through the data-request portal at https://naccdata.org/requesting-data/data-request-process. Version v67 (data freeze June 2024) was used. The neuroimaging linkage files for amyloid and tau PET are distributed by NACC under the same data-use agreement.

Data collection and sharing for the Alzheimer’s Disease Neuroimaging Initiative (ADNI) is funded by the National Institute on Aging (National Institutes of Health Grant U19 AG024904). The grantee organization is the Northern California Institute for Research and Education. In the past, ADNI has also received funding from the National Institute of Biomedical Imaging and Bioengineering, the Canadian Institutes of Health Research, and private sector contributions through the Foundation for the National Institutes of Health (FNIH) including generous contributions from the following: AbbVie, Alzheimer’s Association; Alzheimer’s Drug Discovery Foundation; Araclon Biotech; BioClinica, Inc.; Biogen; Bristol-Myers Squibb Company; CereSpir, Inc.; Cogstate; Eisai Inc.; Elan Pharmaceuticals, Inc.; Eli Lilly and Company; EuroImmun; F. Hoffmann-La Roche Ltd and its affiliated company Genentech, Inc.; Fujirebio; GE Healthcare; IXICO Ltd.; Janssen Alzheimer Immunotherapy Research & Development, LLC.; Johnson & Johnson Pharmaceutical Research &Development LLC.; Lumosity; Lundbeck; Merck & Co., Inc.; Meso Scale Diagnostics, LLC.; NeuroRx Research; Neurotrack Technologies; Novartis Pharmaceuticals Corporation; Pfizer Inc.; Piramal Imaging; Servier; Takeda Pharmaceutical Company; and Transition Therapeutics.

## Code availability

Upon request

## Acknowledgements

The NACC database is funded by NIA/NIH Grant U24 AG072122. NACC data are contributed by the NIA-funded ADRCs: P30 AG062429 (PI James Brewer, MD, PhD), P30 AG066468 (PI Oscar Lopez, MD), P30 AG062421 (PI Bradley Hyman, MD, PhD), P30 AG066509 (PI Thomas Grabowski, MD), P30 AG066514 (PI Mary Sano, PhD), P30 AG066530 (PI Helena Chui, MD), P30 AG066507 (PI Marilyn Albert, PhD), P30 AG066444 (PI John Morris, MD), P30 AG066518 (PI Jeffrey Kaye, MD), P30 AG066512 (PI Thomas Wisniewski, MD), P30 AG066462 (PI Scott Small, MD), P30 AG072979 (PI David Wolk, MD), P30 AG072972 (PI Charles DeCarli, MD), P30 AG072976 (PI Andrew Saykin, PsyD), P30 AG072975 (PI David Bennett, MD), P30 AG072978 (PI Neil Kowall, MD), P30 AG072977 (PI Robert Vassar, PhD), P30 AG066519 (PI Frank LaFerla, PhD), P30 AG062677 (PI Ronald Petersen, MD, PhD), P30 AG079280 (PI Eric Reiman, MD), P30 AG062422 (PI Gil Rabinovici, MD), P30 AG066511 (PI Allan Levey, MD, PhD), P30 AG072946 (PI Linda Van Eldik, PhD), P30 AG062715 (PI Sanjay Asthana, MD, FRCP), P30 AG072973 (PI Russell Swerdlow, MD), P30 AG066506 (PI Todd Golde, MD, PhD), P30 AG066508 (PI Stephen Strittmatter, MD, PhD), P30 AG066515 (PI Victor Henderson, MD, MS), P30 AG072947 (PI Suzanne Craft, PhD), P30 AG072931 (PI Henry Paulson, MD, PhD), P30 AG066546 (PI Sudha Seshadri, MD), P20 AG068024 (PI Erik Roberson, MD, PhD), P20 AG068053 (PI Justin Miller, PhD), P20 AG068077 (PI Gary Rosenberg, MD), P20 AG068082 (PI Angela Jefferson, PhD), P30 AG072958 (PI Heather Whitson, MD), P30 AG072959 (PI James Leverenz, MD).

## Extended Data Tables

**Table 1.**
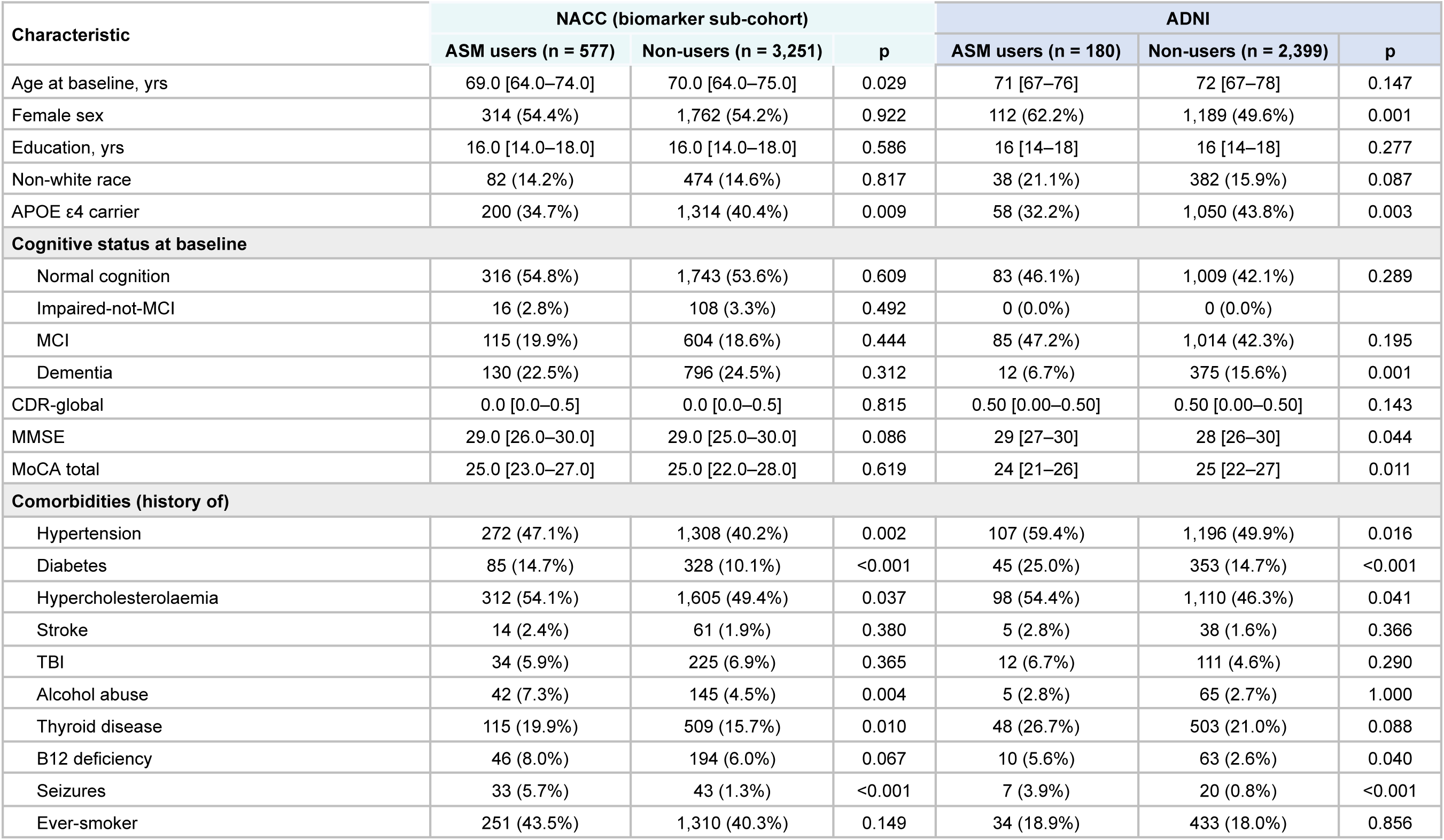
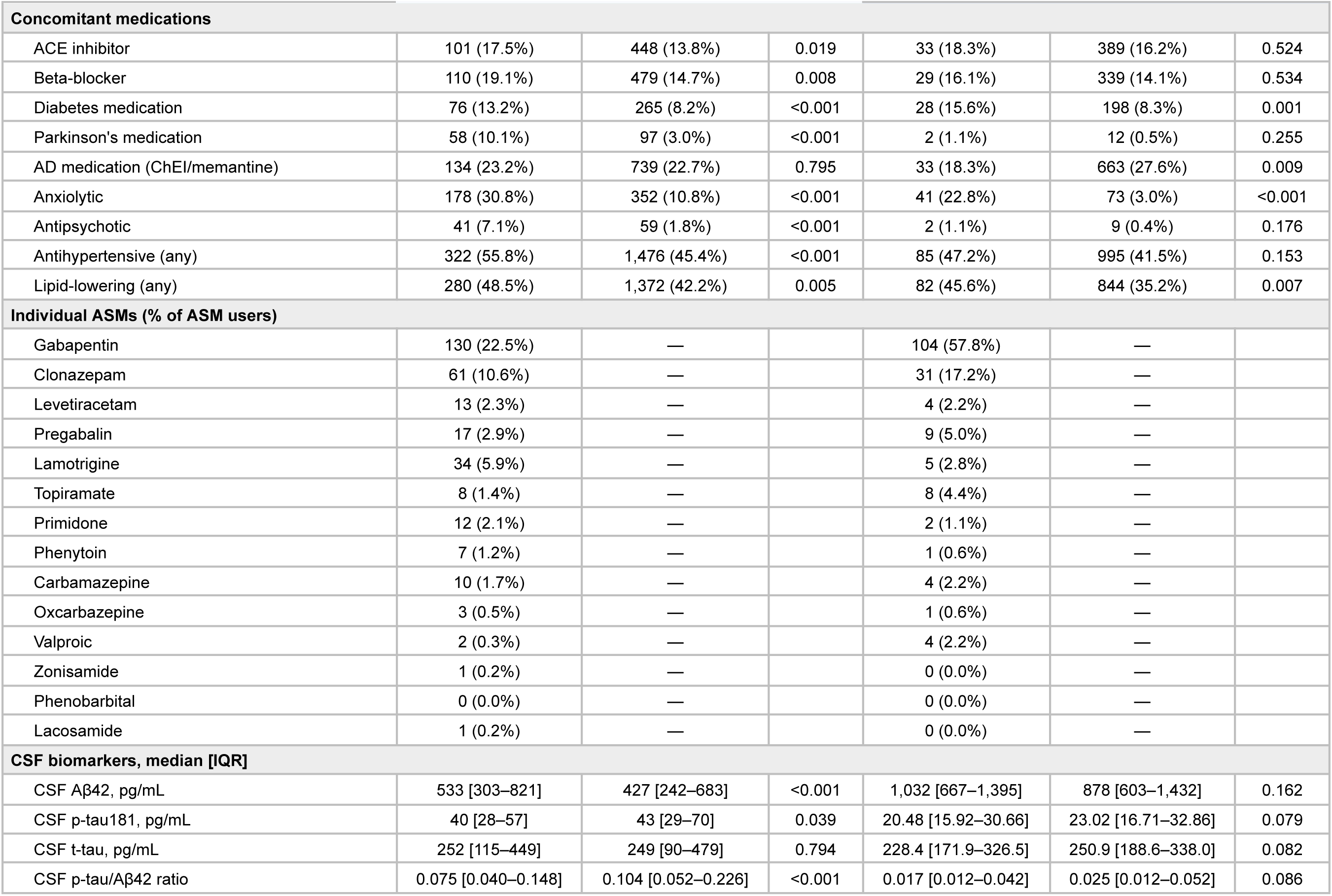
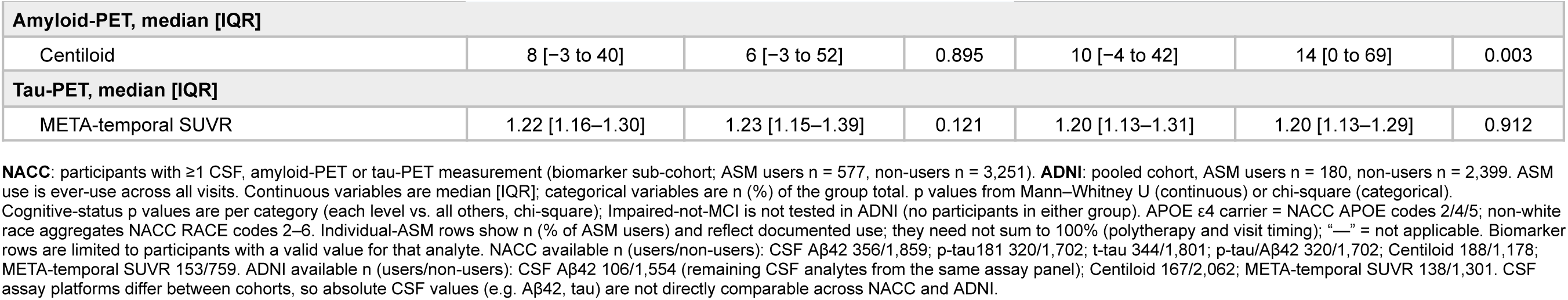
Baseline characteristics of the biomarker sub-cohorts, stratified by antiseizure-medication (ASM) use, in NACC and ADNI.

## Extended Data Figures

**Extended Data Figure 1.**
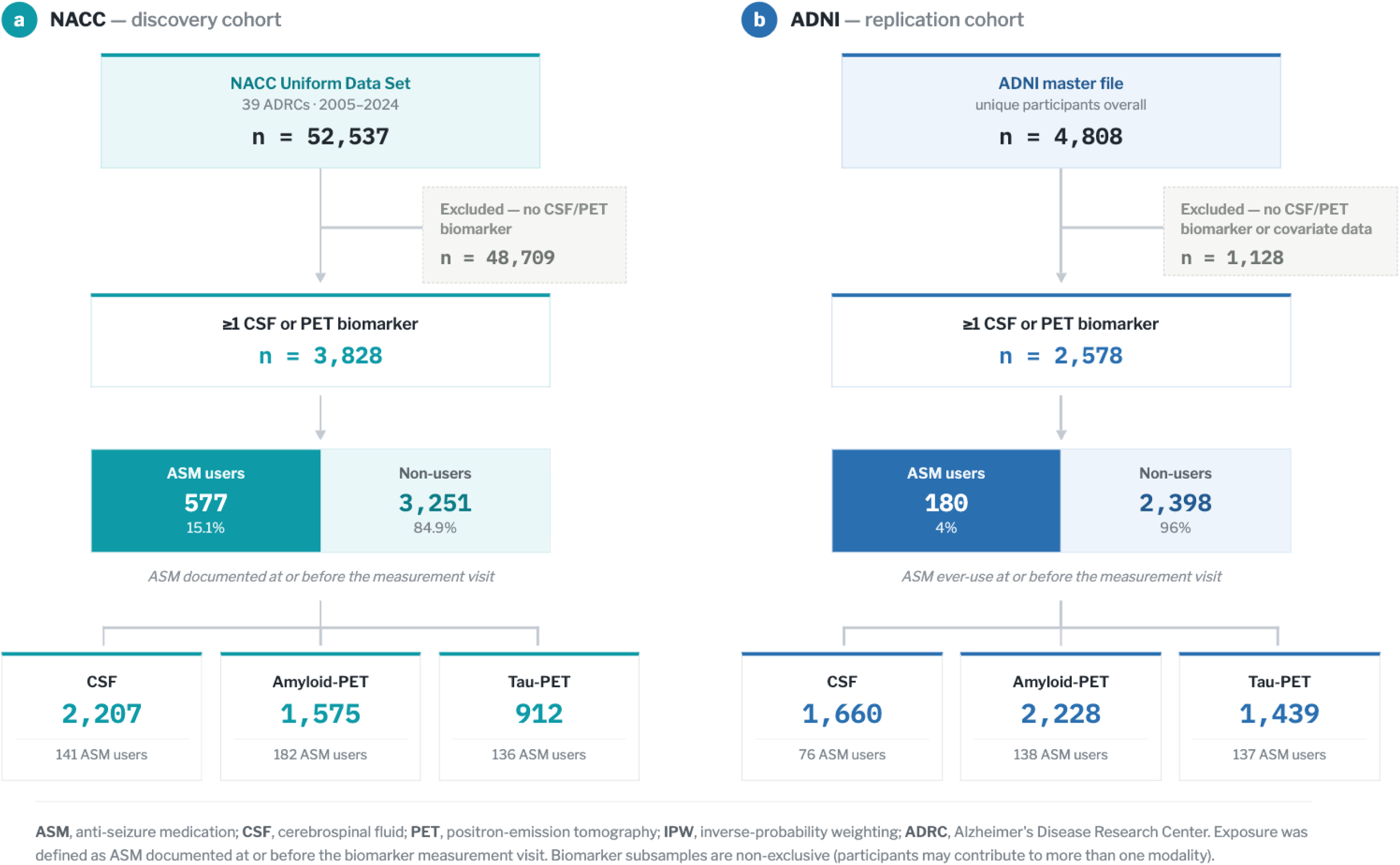
Study flowchart

## References

1. Jack, C. R. et al. NIA-AA Research Framework: Toward a biological definition of Alzheimer’s disease. Alzheimers Dement. 14, 535–562 (2018).

2. Busche, M. A. et al. Critical role of soluble amyloid-β for early hippocampal hyperactivity in a mouse model of Alzheimer’s disease. Proc. Natl. Acad. Sci. 109, 8740–8745 (2012).

3. Minkeviciene, R., Rheims, S., Dobszay, M. B. & al, et. Amyloid β-induced neuronal hyperexcitability triggers progressive epilepsy. J Neurosci 29, 3453–3462 (2009).

4. Zott, B. et al. A vicious cycle of β amyloid–dependent neuronal hyperactivation. Science 365, 559–565 (2019).

5. Rao, N. R. et al. Levetiracetam prevents Aβ production through SV2a-dependent modulation of APP processing in Alzheimer’s disease models. Sci. Transl. Med. 18, eadp3984 (2026).

6. Cirrito, J. R. et al. Synaptic Activity Regulates Interstitial Fluid Amyloid-Beta Levels in Vivo. Neuron 48, 913–922 (2005).

7. Bero, A. W. et al. Neuronal Activity Regulates the Regional Vulnerability to Amyloid-β Deposition. Nat. Neurosci. 14, 750–756 (2011).

8. Busche, M. A. et al. Clusters of Hyperactive Neurons Near Amyloid Plaques in a Mouse Model of Alzheimer’s Disease. Science 321, 1686–1689 (2008).

9. Sanchez, P. E., Zhu, L., Verret, L. & al, et. Levetiracetam suppresses neuronal network dysfunction and reverses synaptic and cognitive deficits in an Alzheimer’s disease model. Proc Natl Acad Sci USA 109, E2895–E2903 (2012).

10. Nygaard, H. B. et al. Brivaracetam, but not ethosuximide, reverses memory impairments in an Alzheimer’s disease mouse model. Alzheimers Res. Ther. 7, 25 (2015).

11. Pooler, A. M., Phillips, E. C., Lau, D. H. W., Noble, W. & Hanger, D. P. Physiological release of endogenous tau is stimulated by neuronal activity. EMBO Rep. 14, 389–394 (2013).

12. Wu, J. W. et al. Neuronal Activity Enhances Tau Propagation and Tau Pathology in Vivo. Nat. Neurosci. 19, 1085–1092 (2016).

13. Yamada, K. et al. Neuronal activity regulates extracellular tau in vivo. J. Exp. Med. 211, 387–393 (2014).

14. Delbono, O., Wang, Z.-M. & Messi, M. L. The rise and deceleration of neuronal excitability in aging and Alzheimer’s disease: Mechanisms, implications, and therapeutic targets. Ageing Res. Rev. 114, 102999 (2026).

15. Targa Dias Anastacio, H., Matosin, N. & Ooi, L. Neuronal hyperexcitability in Alzheimer’s disease: what are the drivers behind this aberrant phenotype? Transl. Psychiatry 12, 257 (2022).

16. Verret, L. et al. Inhibitory Interneuron Deficit Links Altered Network Activity and Cognitive Dysfunction in Alzheimer Model. Cell 149, 708–721 (2012).

17. Vossel, K. A. et al. Incidence and impact of subclinical epileptiform activity in Alzheimer’s disease. Ann. Neurol. 80, 858–870 (2016).

18. Lam, A. D. et al. Association of Seizure Foci and Location of Tau and Amyloid Deposition and Brain Atrophy in Patients With Alzheimer Disease and Seizures. Neurology 103, e209920 (2024).

19. Zhang, M.-Y. et al. Lamotrigine attenuates deficits in synaptic plasticity and accumulation of amyloid plaques in APP/PS1 transgenic mice. Neurobiol. Aging 35, 2713–2725 (2014).

20. Li, L. et al. Autophagy Enhancer Carbamazepine Alleviates Memory Deficits and Cerebral Amyloid-α Pathology in a Mouse Model of Alzheimer’s Disease. Curr. Alzheimer Res. 10, 433–441 (2013).

21. Bakker, A., Krauss, G. L., Albert, M. S. & al, et. Reduction of hippocampal hyperactivity improves cognition in amnestic mild cognitive impairment. Neuron 74, 467–474 (2012).

22. Bakker, A., Rani, N., Mohs, R. & Gallagher, M. The HOPE4MCI study: AGB101 treatment slows progression of entorhinal cortex atrophy in APOE ε4 non-carriers with mild cognitive impairment due to Alzheimer’s disease. Alzheimers Dement. Transl. Res. Clin. Interv. 10, e70004 (2024).

23. Sen, A. et al. Safety, tolerability, and efficacy outcomes of the Investigation of Levetiracetam in Alzheimer’s disease (ILiAD) study: a pilot, double-blind placebo-controlled crossover trial. Epilepsia Open 9, 2353–2364 (2024).

24. Ferreira-Atuesta, C. et al. Sodium channel blockers are associated with reduced dementia risk in late-onset unexplained epilepsy. Preprint at 10.64898/2026.05.20.26353714 (2026).

25. Beekly, D. L. et al. The National Alzheimer’s Coordinating Center (NACC) Database: The Uniform Data Set. Alzheimer Dis. Assoc. Disord. 21, 249–258 (2007).

26. Buckner, R. L. et al. Molecular, Structural, and Functional Characterization of Alzheimer’s Disease: Evidence for a Relationship between Default Activity, Amyloid, and Memory. J. Neurosci. 25, 7709–7717 (2005).

27. Palop, J. J. & Mucke, L. Network abnormalities and interneuron dysfunction in Alzheimer disease. Nat Rev Neurosci 17, 777–792 (2016).

28. Vossel, K. A., Tartaglia, M. C., Nygaard, H. B. & et al. Epileptic activity in Alzheimer’s disease: causes and clinical relevance. Lancet Neurol 16, 311–322 (2017).

29. Tabuena, D. R. et al. Neuronal APOE4-induced early hippocampal network hyperexcitability in Alzheimer’s disease pathogenesis. *Nat*. Aging 6, 886–904 (2026).

30. Andrews-Zwilling, Y. et al. Apolipoprotein E4 Causes Age- and Tau-Dependent Impairment of GABAergic Interneurons, Leading to Learning and Memory Deficits in Mice. J. Neurosci. 30, 13707–13717 (2010).

31. Braak, H. & Braak, E. Neuropathological stageing of Alzheimer-related changes. Acta Neuropathol. (Berl.) 82, 239–259 (1991).

32. Fischl, B. FreeSurfer. NeuroImage 62, 774–781 (2012).

